# A mathematical model to estimate percentage secondary infections from margin of error of diagnostic sensitivity: Useful tool for regulatory agencies to assess the risk of propagation due to false negative outcome of diagnostics

**DOI:** 10.1101/2021.01.29.21250804

**Authors:** P Azhahianambi, K Karthik, RP Aravindh Babu, TMA Senthil kumar, C Balachandran, G Dhinakar Raj

## Abstract

False negative outcome of a diagnosis is one the major reasons for the dissemination of the diseases with high risk of propagation. Diagnostic sensitivity and the margin of error determine the false negative outcome of the diagnosis. A mathematical model had been developed to estimate the mean % secondary infections based on the margin of error of diagnostic sensitivity, % prevalence and R_0_ value. This model recommends a diagnostic test with diagnostic sensitivity ≥ 96% and at least 92% lower bound limit of the 95% CI or ≤ 4% margin of error for a highly infectious diseases like COVID-19 to curb the secondary transmission of the infection due to false negative cases. Positive relationship was found between mean % secondary infection and margin of error of sensitivity suggesting greater the margin of error of a diagnostic test sensitivity, higher the number of secondary infections in a population due to false negative cases. Negative correlation was found between number of COVID-19 test kits (>90% sensitivity) with regulatory approval and margin of error (R= −0.92, *p*=0.023) suggesting lesser the margin of error of a diagnostic test, higher the chances of getting approved by the regulatory agencies. However, there are no specific regulatory standards available for margin of error of the diagnostic sensitivity of COVID-19 diagnostic tests. Highly infectious disease such as COVID-19, certainly need specific regulatory standards on margin of error or 95% CI of the diagnostic sensitivity to curb the dissemination of the disease due to false negative cases and our model can be used to set the standards such as sensitivity, margin of error or lower bound limit of 95% CI.

## 1. Introduction

A perfect diagnostic test does not exist in real life and therefore diagnostic procedures can make only partial distinction between subjects with and without disease (Simundic, 2009). Diagnostic sensitivity and specificity of a diagnostic test are the metrices generally considered to assess the performance of the test in the real world or clinical condition. Sensitivity defines the proportion of true positive subjects with the disease in a total group of subjects with the disease and specificity defines proportion of subjects without the disease with negative test result in total of subjects without disease (Simundic, 2009). Among the diagnostic sensitivity and specificity, diagnostic sensitivity of a test is very critical in diagnosis of infectious diseases with high risk of propagation as false negative results could facilitate the community transmission of infectious diseases such as COVID-19. The precision of diagnostic sensitivity is also an important factor in the diagnosis of highly infectious diseases which is determined by 95% confidence interval (CI). Narrower the 95% CI interval, higher the precision of the diagnostic test. An ideal diagnostic test would have highest sensitivity and specificity with very narrow 95% CI which hardly exist in the real world. However, a test with diagnostic sensitivity close to 100% and very narrow 95% CI is need of the hour for the infectious disease such as COVID-19 with basic reproduction number (R_0_) greater than one. R_0_ is defined as the expected number of secondary infections produced by a single infection in a completely susceptible population and If the mean R_0_ > 1, the infection will spread exponentially. R_0_ estimates for SARS virus infections have been reported to range between 2 and 5. The R_0_ value of SARS-CoV-2 infection during the early period of outbreak in China was estimated to be about 2.0 (Liu et al., 2020a) and the World Health Organization (WHO) estimate of R_0_ value was 1.95 (1.4-2.5) (WHO, 2020). The recent studies claimed that the mean R_0_ value of COVID-19 disease had been estimated to be 3.28 (Liu et al., 2020b) and the recently mutated SARS-CoV-2 B.1.1.7 lineage has R_0_ value higher than the previous lineages. Higher the diagnostic sensitivity and narrower the 95% CI together churn out less false negative results and help in curbing the community transmission of highly infectious diseases with pandemic potential. A 95% CI is a range of values that can be 95% certain contains the true mean of the population and one can be 95% confident that the true population parameter is between the lower and upper limit values. US-FDA recommended that diagnostic sensitivity and specificity of all the tests to be reported with 95 % CI (FDA, 2011; Hess et al., 2012). The confidence interval range of a diagnostic test can be simply summarised in a number as Margin of Error (M) which will help the clinicians to easily understand the precision of the diagnostic test. The margin of error is simply one-half the width of the confidence interval, i.e. (Upper limit − Lower limit)/2 (Hess et al., 2012). Lower the margin of error of the diagnostic sensitivity and specificity, higher the precision of the diagnostic test. There is no commonly accepted standard for how small the margin of error should be for diagnostic test. In most of the social science studies a margin of error < 5% is the acceptable limit, but no such published or regulatory guidelines exists for the diagnostic tests used in the veterinary and medical practice (Royse, 2008; Sullivan, 2008; Royse et al., 2010). Though smaller margin of error are preferred for any diagnostic test, diagnostic test intended for the highly infectious diseases with R_0_>1 needs specific recommendation from the regulatory agencies to restrict the rapid spread of the disease. Sensitivity and specificity of a diagnostic test may vary with the prevalence of disease in a population in which higher the prevalence of disease, higher the sensitivity and lower the specificity of the diagnostic test (Leeflang et al., 2013). A mathematical model had been developed to estimate the mean % secondary infections based on the margin of error of diagnostic sensitivity, % prevalence and R_0_ value which can be used to set norms for diagnostic tests intended for diseases with high risk of propagation. This paper also discusses the role of diagnostic sensitivity and margin of error in the risk of propagation of the diseases with R_0_ > 1 and emphasis the need to consider the margin of error of a diagnostic sensitivity of a diagnostic test as one of the parameters while approving the diagnostic tests associated with high risk of disease propagation.

## 2. Materials and Methods

### 2.1. Mathematical model to estimate the mean secondary infection in relation to diagnostic sensitivity and margin of error

The false negative cases are the persons carrying the infection but failed to be detected by the diagnostic test. Unlike false positive cases, the false negative cases carrying highly infectious pathogens such as COVID-19 virus could potentially act as community spreaders and source of secondary infections (**Fig.1**). If the R_0_ value is 2.0, every false negative person can potentially transmit the virus to 2 persons in a population. The mean % secondary infections related with the % false negative cases were estimated using a customised formula. The range of sensitivity in increment of one corresponding to the margin of error and length of range were kept as variable. The R_0_ value and % prevalence values were kept as constants. The R_0_ value was kept as 2.0 in this formula as R_0_ value of COVID-19 (WHO, 2020; Liu et al 2020a). Percentage prevalence of COVID-19 was assigned as 10% (0.10) as the mean weekly COVID-19 positive in USA was 9.6% (5%-22%) (CDC, 2020). Mean % secondary infection due to false negative results was estimated for the diagnostic tests with sensitivity of 100%, 99%,98%,97%,96%,95% and 90% with margin of error ranging from 1% to 10%. The margin of error (*M*) was calculated using the formula, *Upper limit of 95% CI − Lower limit of 95% CI)/2* (Hess et al., 2012). The mean % secondary infection for a diagnostic test with a sensitivity (i.e., 95%) and margin of error (i.e., 2%) was calculated as per the following formula.

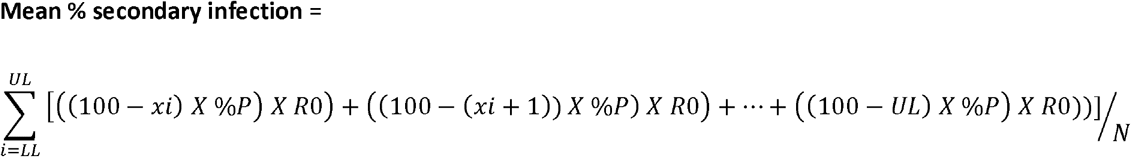

**x = 95% CI range**

***i*= Lower limit of the 95% CI**

**UL= Upper limit of the 95% CI**

**%P= Percentage prevalence of COVID-19 (Assigned as 10% prevalence (0.10), Mean weekly COVID-19 positive in USA 9.6% (5%-22%))**

**R**_**0**_ **= Basic reproduction number (Assigned as constant value 2.0, R0 value of SARS-CoV-2 infection)**

**N= Number of random populations**

For easy calculation of the mean % secondary infection for a diagnostic test with any range of sensitivity, variable R_0_ and % prevalence, a R-software programme had been scripted based on the aforementioned formula.

**Fig 1:**
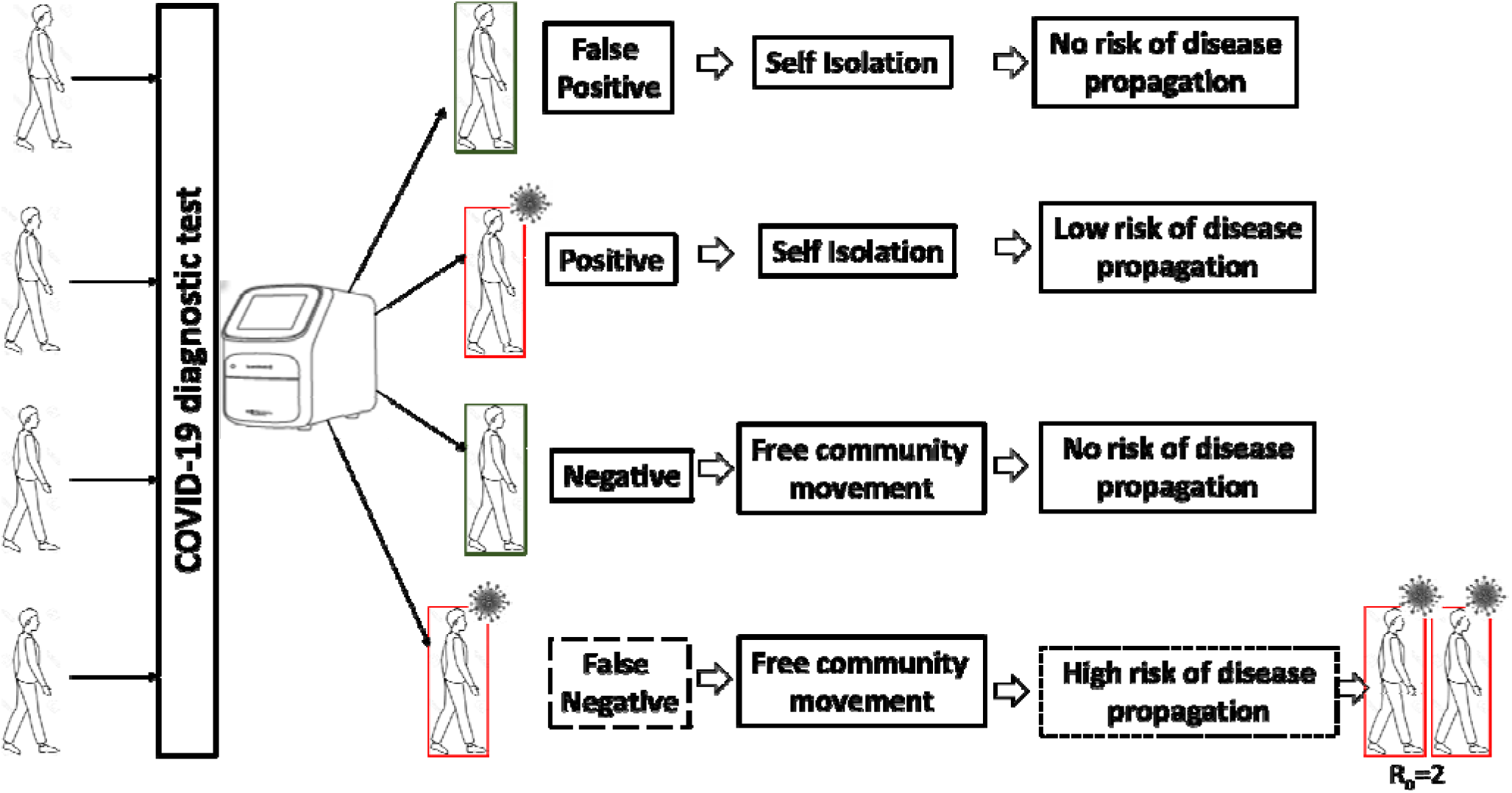
Diagram explaining the risk of propagation of highly infectious diseases such as COVID-19 due to false negative outcome of diagnosis

The R software programme to calculate the secondary infection for a margin of error. Here the variables are ‘x’ and ‘n’; the constants are ‘y’ and ‘z’.

**x = 95% CI as i.e**., **c (96:100)**

**n= number of random populations**

**y= Percentage prevalence of COVID-19 (0.1)**

**z= R**_**0**_ **value (2)**

**h= mean % secondary infection**

**++++++START OF FUNCTION++++++++**

infection <-function (x,y,z)

{

n = length (x) x1 = 0

for (i in 1:n)

{

x1[i] = 100-x[i]

}

g = sum (x1*y*z) h = g/n

cat (“The value of g and h are”, g, “and”, h, “respectively”, “\n”)

}

**++++++END OF FUNCTION++++++++**

The range of 95% CI was taken as simple number range (i.e., 94%-96%) where the lower bound and upper bound limits are 94% and 96%, respectively. The % secondary infections were calculated for the given 95% CI (94%-96%) by subtracting the lower limit value (x_i_=94%) from 100% sensitivity and multiplied by % prevalence (0.10) and the product finally multiplied by R_0_ value [(100-94) X 0.10 × 2=1.2]. Similarly, the % secondary infections were calculated for 95% (x_i_+1) and 96% (UL). The mean % secondary infection was estimated as the mean of the 3 values. The diagnostic test with 95% diagnostic sensitivity (95% CI, 94%-96%) is expected to have the population sensitivity from 94% to 96% with a confidence of 95%. Here in the model, in a random of three infected populations with the disease prevalence of 10%, the diagnostic test has been assumed to have 94%, 95% and 96% diagnostic sensitivity, respectively. The mean % secondary infection from the false negative cases of the diagnostic test with 95% sensitivity (95% CI, 94%-96%) in three random populations is estimated to be 1.0 [(1.2+1.0+0.8)/3]. This model assumes that a diagnostic test with 95% diagnostic sensitivity with margin of error 1 (95% CI, 94%-96%) could facilitate the spread of an infectious disease (R_0_=2.0) up to 1% of the population as secondary infection due to false negative results. A diagnostic test with low false negative outcome which could spread an infectious disease such as COVID-19 to < 1% of the population is an ideal one.

### 2.2. COVID-19 diagnostics data

The data on antibody, antigen and Nucleic Acid Amplification Test (NAAT) based COVID-19 diagnostic kits, diagnostic sensitivity and specificity of the kits, margin of error and regulatory approval status of the COVID-19 diagnostic kits were obtained from *finddx*.*com*. The total number of antibody, antigen and NAAT based COVID-19 diagnostic kits with complete data set, kits with diagnostic sensitivity and specificity greater than 90%, margin of error of the diagnostic kits and list of COVID-19 diagnostic kits with regulatory approvals were obtained from *finddx*.*com* on 30^th^ November 2020. The antibody and NAAT based COVID-19 diagnostic test kits with both diagnostic sensitivity and specificity greater than 90% were categorized based on the margin of error values.

### 2.3. Statistical analysis

The data generated from the model and the COVID-19 diagnostics data obtained from *finddx*.*com* were analysed by R-software R-4.0.2, GraphPad prism 7.0 and MS excel. Pearson correlation coefficient and coefficient of determination between the margin of error and mean % secondary infections were calculated and line plot was plotted. Pearson correlation coefficient and coefficient of determination between the margin of error of diagnostic sensitivity and specificity and number of COVID-19 diagnostic kits approved by regulatory agencies with varying margin of error were calculated. Linear regression analysis of interval-valued data such as range of margin or error and number of approved kits in each range of margin of error was calculated using the upper, lower and midpoint values of the margin of error range (de Carvalho, et al., 2004)

## 3. Results

### 3.1. Positive Correlation between margin of error of diagnostic test sensitivity and mean % secondary infection

The mean % secondary infections were estimated for the diagnostic tests with sensitivity of 100%, 99%, 98%, 97%, 96%, 95% and 90% with margin of error ranging from 1% to 10%. The secondary infections are assumed to be due to the false negative cases spreading the infections in a community where there is no herd immunity to the disease. The model suggests that diagnostic tests with sensitivity ≥ 96% with margin of error ≤ 4, could be an ideal test with low false negative outcome that could spread the disease with R_0_ value 2 and 10% prevalence to less than 1% of the population as secondary infection (**Table. 1**). A diagnostic test with risk of propagation of disease to less than 1% of population is expected to curb the spread of disease. Hence, less than 1% mean secondary infection was considered as the cut-off value to determine the diagnostic sensitivity, 95% CI and margin of error. The R software programme can be used to calculate the mean % secondary infection for any variable diagnostic sensitivity range, R_0_ values and % prevalence ranges.

**Table 1.**
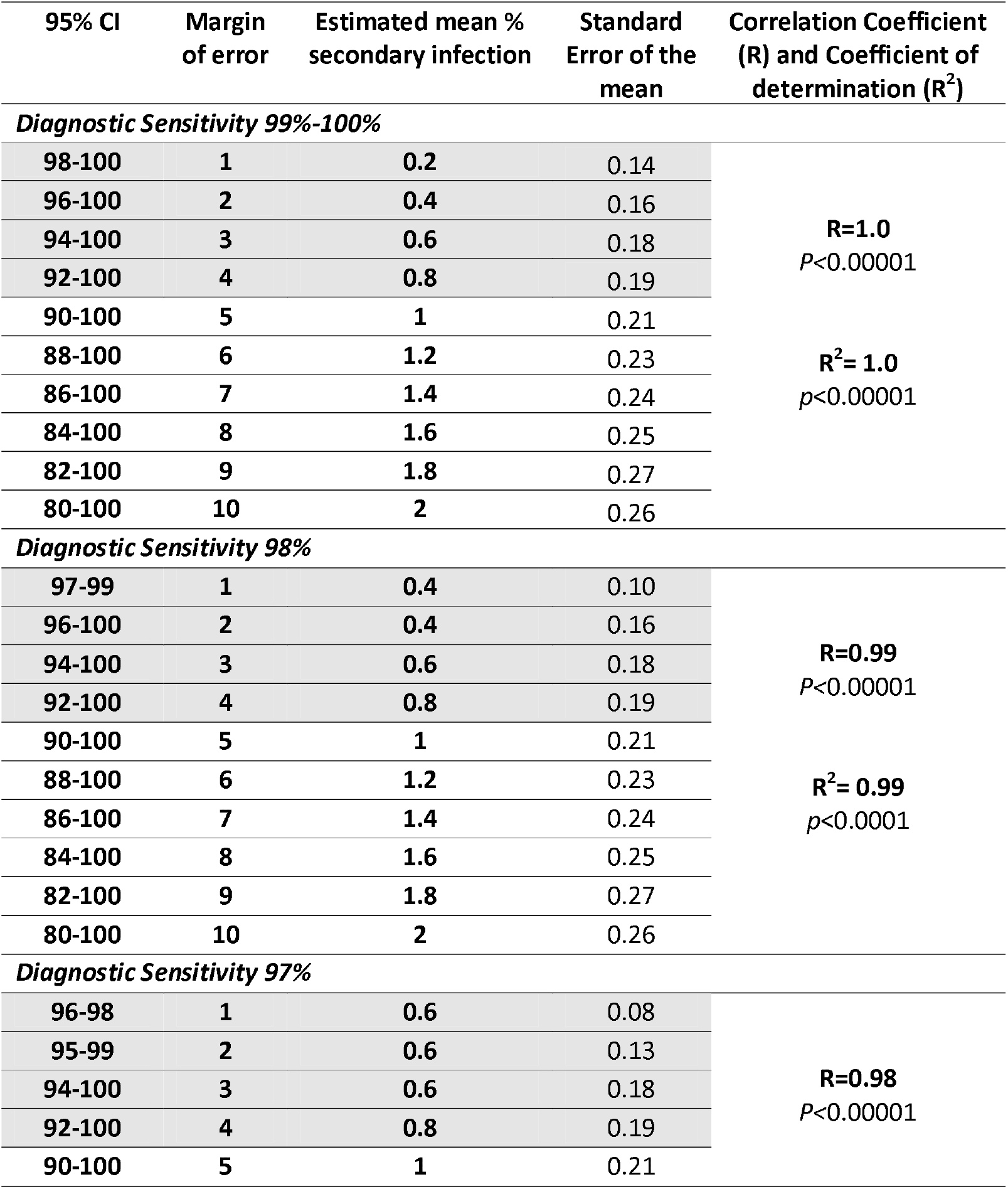
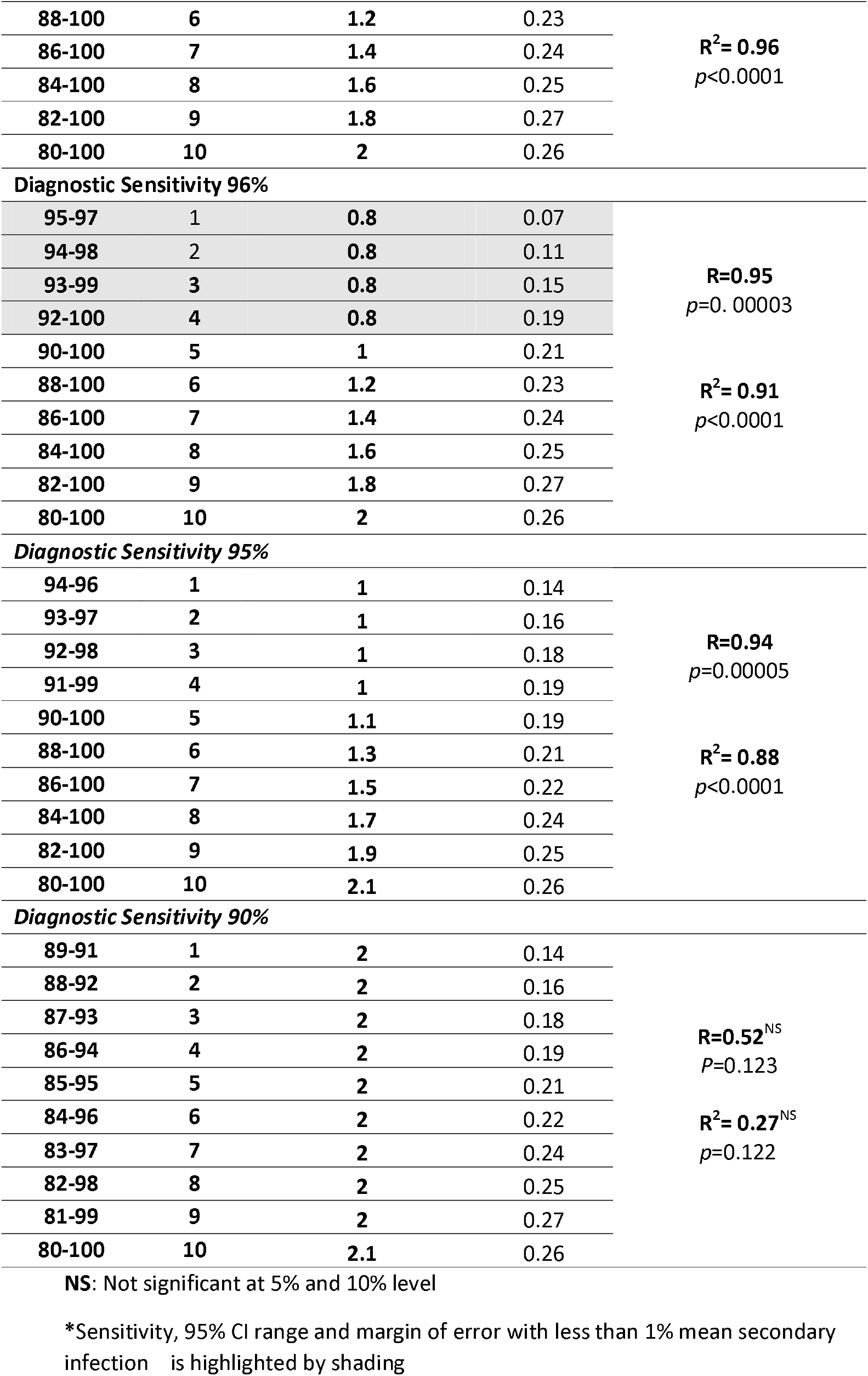
Estimated mean % secondary infection at various diagnostic sensitivity and margin of error values where R_0_ is 2.0 and relationship between margin of error and estimated mean % secondary infection

| 95% CI | Margin of error | Estimated mean % secondary infection | Standard Error of the mean | Correlation Coefficient (R) and Coefficient of determination (R<sup>2</sup>) |
| --- | --- | --- | --- | --- |
| Diagnostic Sensitivity 99%-100% |  |  |  |  |
| 98-100 | 1 | 0.2 | 0.14 | R=1.0<br>P<0.00001 |
| 96-100 | 2 | 0.4 | 0.16 |  |
| 94-100 | 3 | 0.6 | 0.18 |  |
| 92-100 | 4 | 0.8 | 0.19 |  |
| 90-100 | 5 | 1 | 0.21 |  |
| 88-100 | 6 | 1.2 | 0.23 | R<sup>2</sup>= 1.0<br>p<0.00001 |
| 86-100 | 7 | 1.4 | 0.24 |  |
| 84-100 | 8 | 1.6 | 0.25 |  |
| 82-100 | 9 | 1.8 | 0.27 |  |
| 80-100 | 10 | 2 | 0.26 |  |
| Diagnostic Sensitivity 98% |  |  |  |  |
| 97-99 | 1 | 0.4 | 0.10 | R=0.99<br>P<0.00001 |
| 96-100 | 2 | 0.4 | 0.16 |  |
| 94-100 | 3 | 0.6 | 0.18 |  |
| 92-100 | 4 | 0.8 | 0.19 |  |
| 90-100 | 5 | 1 | 0.21 |  |
| 88-100 | 6 | 1.2 | 0.23 | R<sup>2</sup>= 0.99<br>p<0.0001 |
| 86-100 | 7 | 1.4 | 0.24 |  |
| 84-100 | 8 | 1.6 | 0.25 |  |
| 82-100 | 9 | 1.8 | 0.27 |  |
| 80-100 | 10 | 2 | 0.26 |  |
| Diagnostic Sensitivity 97% |  |  |  |  |
| 96-98 | 1 | 0.6 | 0.08 | R=0.98<br>P<0.00001 |
| 95-99 | 2 | 0.6 | 0.13 |  |
| 94-100 | 3 | 0.6 | 0.18 |  |
| 92-100 | 4 | 0.8 | 0.19 |  |
| 90-100 | 5 | 1 | 0.21 |  |
| 88-100 | 6 | 1.2 | 0.23 | **R<sup>2</sup>= 0.96**<br>*p*<0.0001 |
| 86-100 | 7 | 1.4 | 0.24 |  |
| 84-100 | 8 | 1.6 | 0.25 |  |
| 82-100 | 9 | 1.8 | 0.27 |  |
| 80-100 | 10 | 2 | 0.26 |  |
| Diagnostic Sensitivity 96% |  |  |  |  |
| 95-97 | 1 | 0.8 | 0.07 | **R=0.95**<br>*p*=0. 00003 |
| 94-98 | 2 | 0.8 | 0.11 |  |
| 93-99 | 3 | 0.8 | 0.15 |  |
| 92-100 | 4 | 0.8 | 0.19 |  |
| 90-100 | 5 | 1 | 0.21 |  |
| 88-100 | 6 | 1.2 | 0.23 | **R<sup>2</sup>= 0.91**<br>*p*<0.0001 |
| 86-100 | 7 | 1.4 | 0.24 |  |
| 84-100 | 8 | 1.6 | 0.25 |  |
| 82-100 | 9 | 1.8 | 0.27 |  |
| 80-100 | 10 | 2 | 0.26 |  |
| Diagnostic Sensitivity 95% |  |  |  |  |
| 94-96 | 1 | 1 | 0.14 | **R=0.94**<br>*p*=0.00005 |
| 93-97 | 2 | 1 | 0.16 |  |
| 92-98 | 3 | 1 | 0.18 |  |
| 91-99 | 4 | 1 | 0.19 |  |
| 90-100 | 5 | 1.1 | 0.19 |  |
| 88-100 | 6 | 1.3 | 0.21 | **R<sup>2</sup>= 0.88**<br>*p*<0.0001 |
| 86-100 | 7 | 1.5 | 0.22 |  |
| 84-100 | 8 | 1.7 | 0.24 |  |
| 82-100 | 9 | 1.9 | 0.25 |  |
| 80-100 | 10 | 2.1 | 0.26 |  |
| Diagnostic Sensitivity 90% |  |  |  |  |
| 89-91 | 1 | 2 | 0.14 | **R=0.52<sup>NS</sup>**<br>*P*=0.123 |
| 88-92 | 2 | 2 | 0.16 |  |
| 87-93 | 3 | 2 | 0.18 |  |
| 86-94 | 4 | 2 | 0.19 |  |
| 85-95 | 5 | 2 | 0.21 |  |
| 84-96 | 6 | 2 | 0.22 | **R<sup>2</sup>= 0.27<sup>NS</sup>**<br>*p*=0.122 |
| 83-97 | 7 | 2 | 0.24 |  |
| 82-98 | 8 | 2 | 0.25 |  |
| 81-99 | 9 | 2 | 0.27 |  |
| 80-100 | 10 | 2.1 | 0.26 |  |

NS: Not significant at 5% and 10% level
\*Sensitivity, 95% CI range and margin of error with less than 1% mean secondary infection is highlighted by shading

Positive correlation was found between mean % secondary infection and margin of error (R=1.0, p=0.0001 (sensitivity 100%); R=0.94, p=0.00005 (sensitivity 95%)) suggesting greater the margin of error of a diagnostic test sensitivity, higher the number of secondary infections in a population due to false negative cases (**Table. 1**) (**Fig.2**).

**Fig 2:**
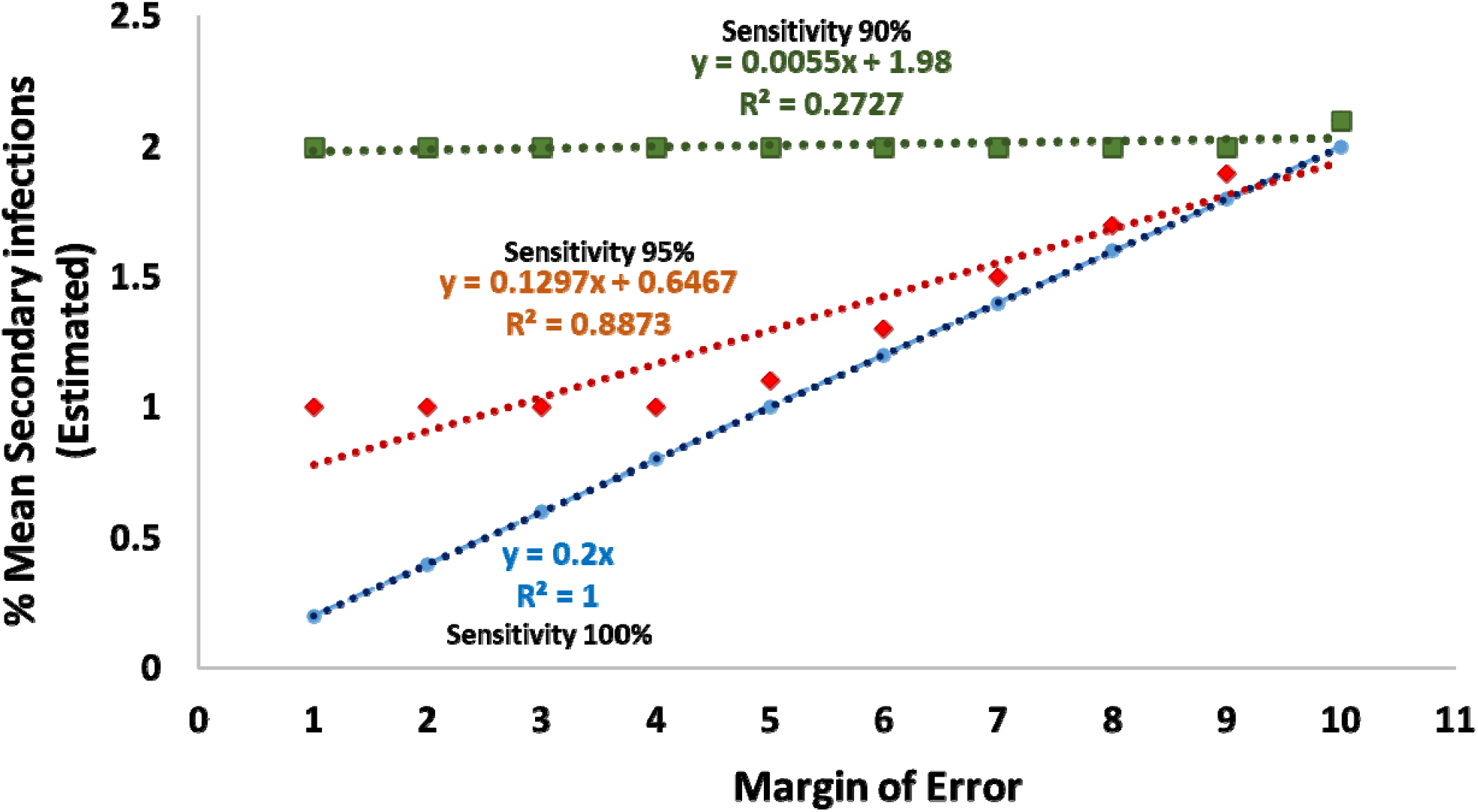
Relationship between the estimated % mean secondary infections and margin of error where % prevalence of the disease is 10% and R_0_ is 2.0

This model recommends a diagnostic test with diagnostic sensitivity ≥ 96% and at least 92% lower bound limit of the 95% CI or ≤ 4% margin of error for highly infectious diseases like COVID-19 to curb the secondary transmission of the infection due to false negative cases. (**Fig. 3**). The recommended sensitivity and margin of error of a diagnostic test will change in accordance with the R_0_ value and % prevalence. Higher the R_0_ value and % prevalence of the disease, we need to have a diagnostic test with higher sensitivity and narrow 95% CI or lower margin of error. High reproduction rate number and prevalence of an infectious disease are expected to drive the spread the disease to a greater number of people in a population and need a diagnostic test with highest sensitivity and lowest margin of error to prevent the secondary spread of the disease due to false negative outcome.

**Fig 3:**
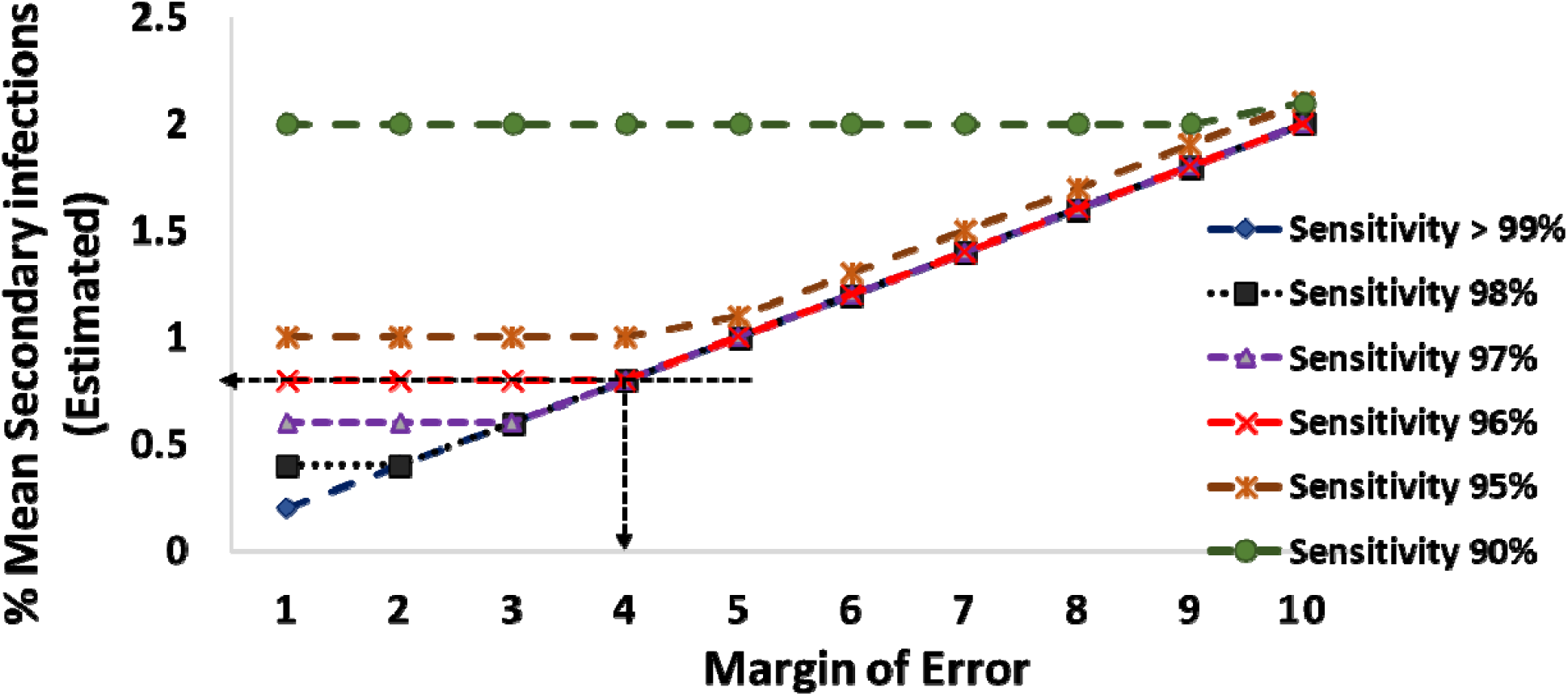
Estimated % mean secondary infections in different diagnostic sensitivity and margin of error where % prevalence of the disease is 10% and R_0_ is 2.0

### 3.2. Approved COVID-19 test kits with margin of error data

Among 73,131 and 6 number of NAAT based, antibody based and antigen based COVID-19 diagnostic kits, respectively listed in the *finddx*.*com*, only 41, 32 and 5 number of kits, respectively were available with complete data set including the 95% CI of sensitivity and specificity. NAAT based, antibody based and antigen based COVID-19 diagnostic kits with diagnostic sensitivity and specificity greater than 90% were 29, 13 and 1, respectively (**Table.2**).

**Table 2.**
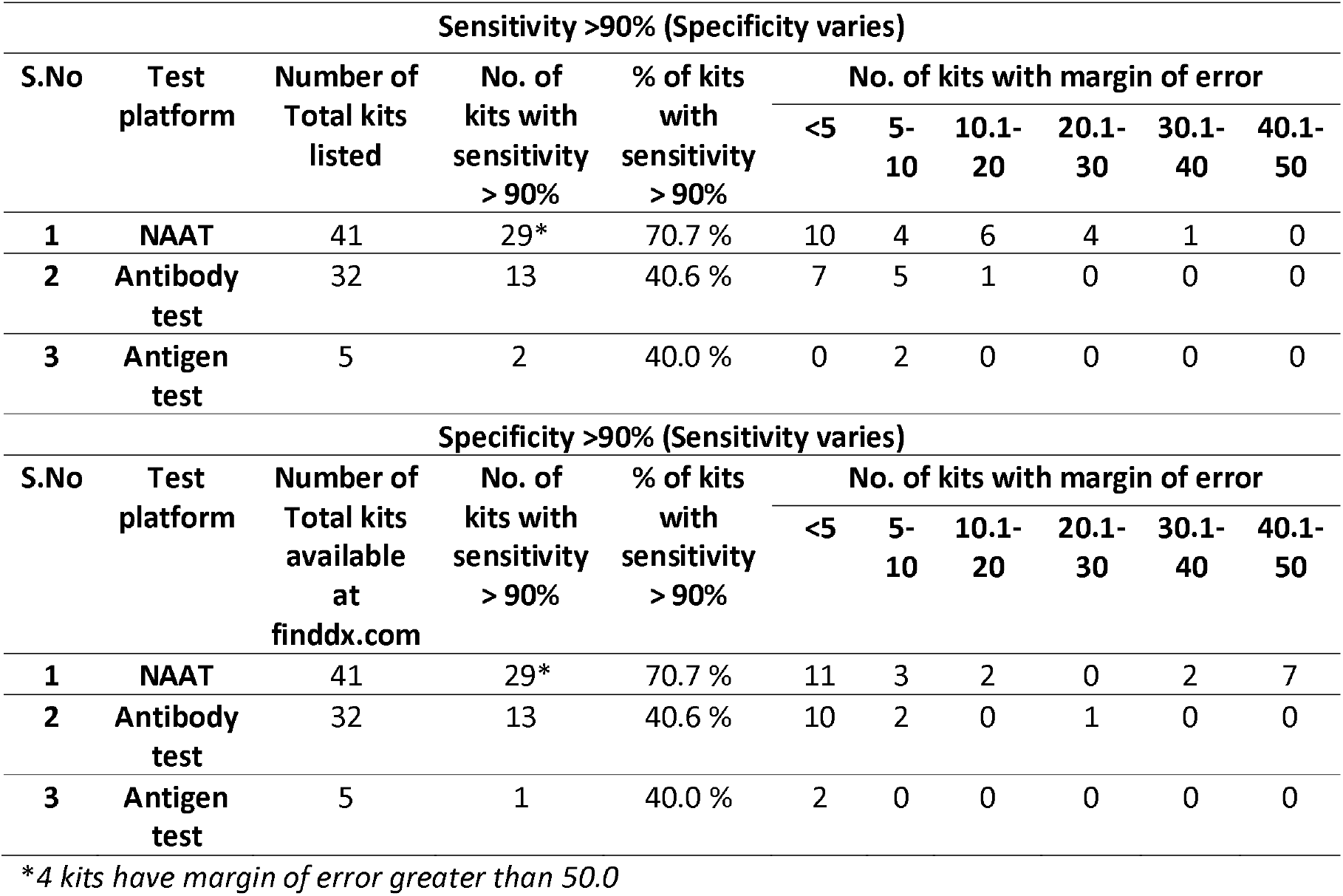
Approved COVID-19 diagnostic kits with diagnostic sensitivity and specificity greater than 90% listed in finddx.com

The number of COVID-19 test kits with diagnostic sensitivity and specificity greater than 90% with varying margin of error were binned into 6 different margin of error intervals (1%-50%). The number of COVID-19 test kits falling under different range of margin error were listed in **Table.2**. The number of NAAT based, antibody based and antigen based COVID-19 diagnostic kits with less than 5% margin of error and >90% sensitivity was 10, 7 and 0, respectively. The number of NAAT based, antibody based and antigen based COVID-19 diagnostic kits with less than 5% margin of error and >90% specificity was 11, 10 and 2, respectively (**Table. 2**).

### 3.3. Negative correlation between the margin of error of diagnostic sensitivity and regulatory approval of COVID-19 test kits

The number of approved NAAT and antibody based COVID-19 diagnostic kits with >90% sensitivity and specificity binned into 6 different margin of error intervals, were subjected to linear regression analysis. Negative correlation was found between number of COVID-19 test kits (>90% sensitivity) with regulatory approval and margin of error (R= −0.92, *p*=0.023) suggesting lesser the margin of error of a diagnostic test, higher the chances of getting approved by the regulatory agencies (**Table. 3**) (**Fig.4**). The margin of error explained 85% variation in the regulatory approval process of the COVID-19 test kits (R^2^=0.85, *p*=0.023) suggesting the regulatory agencies such as US-FDA, CE-mark (EU), Health Canada, Australia TGA and Singapore HAS and Brazil ANVISA considering margin of error or 95% CI while approving the COVID-19 test kits. Though negative correlation was found between number of COVID-19 test kits (>90% specificity) with regulatory approval and margin of error (R= - 0.70, *p*=0.188), it was not statistically significant (**Table. 3**) (**Fig.4**).

**Table 3.**
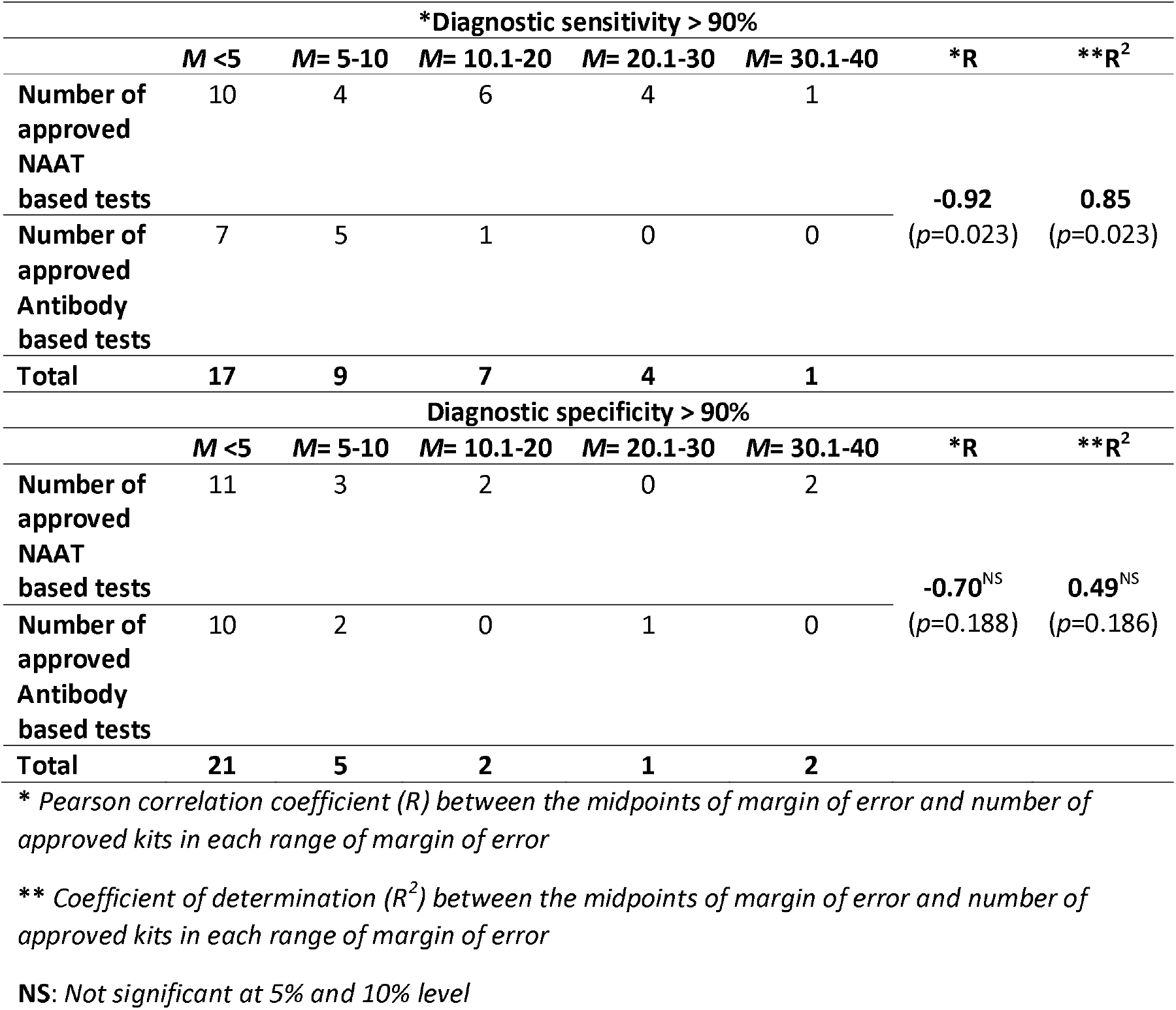
Margin of error of NAAT based and antibody-based COVID-19 test kits with regulatory approval and relationship between margin of error and number of regulatory approved COVID-19 test kits

**Fig 4:**
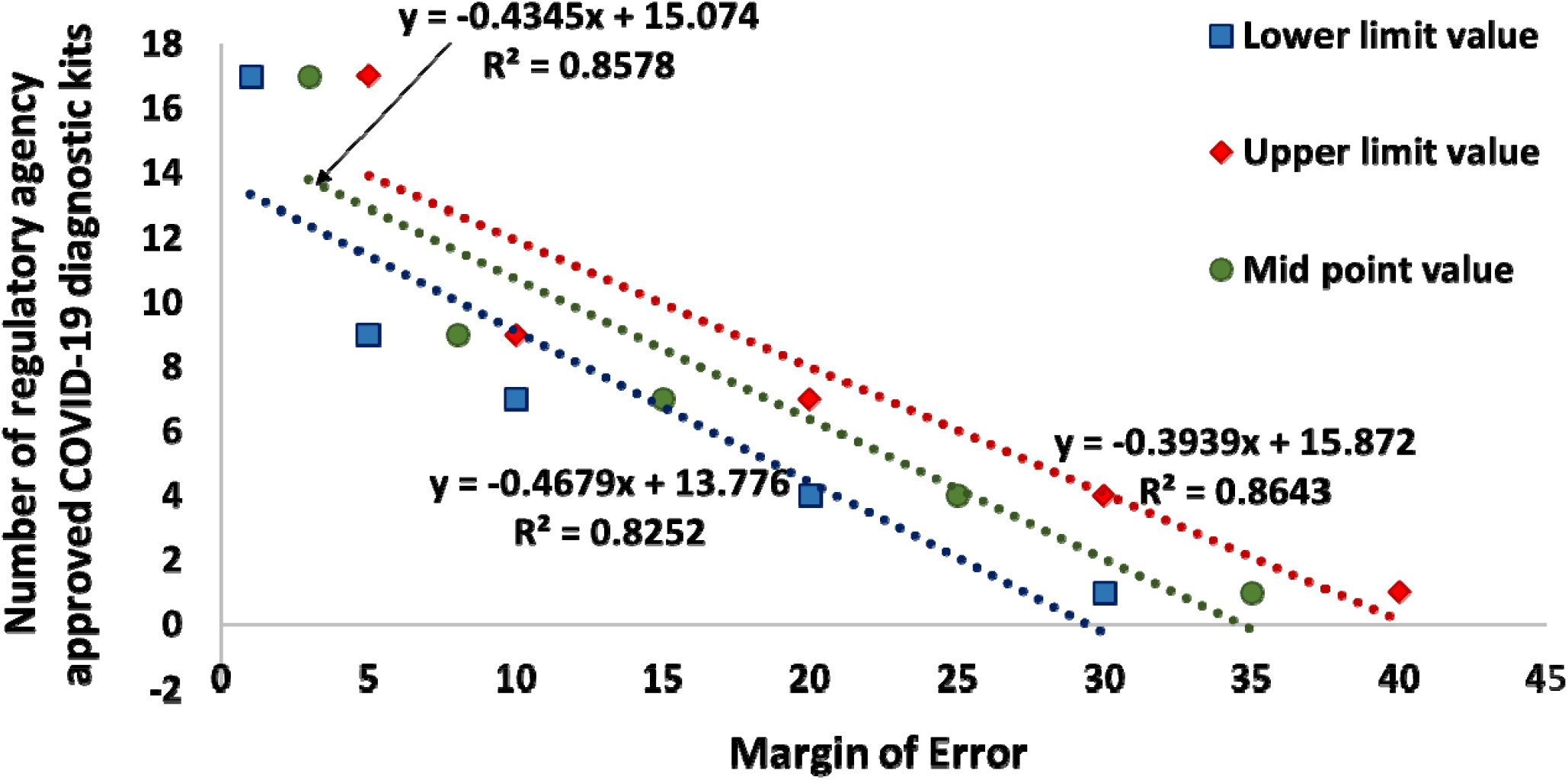
Relationship between the number of COVID-19 diagnostics kits approved by regulatory agencies and margin of error

## 4. Discussion

Diagnostic tests for highly infectious diseases with low sensitivity and wider 95% CI or high margin of error are expected to yield more false negative results which could lead to rapid spread of the infections in community. A systematic review of the accuracy of COVID-19 tests reported false negative rates of between 2% and 29% (equating to sensitivity of 71-98%), based on negative RT-PCR tests which were positive on repeat testing (Arevalo-Rodriguez et al., 2020). A person diagnosed negative for the infectious disease but actually carrying the infectious agent moving freely in the population and allowed to fly all over the world thus unwittingly transmitting the disease agent. Individuals with false negative results tend to relax physical distancing, wearing PPE and other personal measures designed to reduce the transmission of the virus to others (West et al., 2020) thus facilitate dissemination of the disease. Even diagnostic tests with perfect sensitivity but wider 95% CI or high margin of error could yield a greater number of false negative outcomes and that would facilitate the community transmission of the disease. Thus, margin of error of the diagnostic sensitivity of the test is an important parameter as the diagnostic sensitivity of the test.

Though it is well known that the person with false negative test result could act as a silent carrier of the infection and spread the infection, there are no study or model to show the relationship between margin of error of diagnostic sensitivity which led to false negative outcome and secondary spread of the infection. Our model suggests the importance of margin of error of the diagnostic sensitivity in spreading the diseases through the false negative outcome which needs to be validated in field condition.

Though there are regulatory standards on 95% CI or margin of error for the influenza virus diagnosis, currently no regulatory standards on 95% CI or margin of error for the diagnostic sensitivity of COVID-19 diagnostics. FDA considers the observed sensitivity and specificity and their respective 95% lower confidence bounds as part of device performance. As per FDA regulations, the sensitivity for the device testing for influenza A virus must be at least 90 % with a lower bound of the 95 % CI greater than or equal to 80%. The sensitivity for the device testing for influenza B virus must be at least 80 % with a lower bound of the 95 % CI greater than or equal to 70 percent (FDA, 2020). The model used by FDA to estimate the recommended sensitivity and 95% CI is not known. Our model recommends a diagnostic test with diagnostic sensitivity ≥ 96% and at least 92% lower bound limit of the 95% CI or ≤ 4% margin of error for highly infectious diseases like COVID-19 to curb the secondary transmission of the infection due to false negative cases. This recommendation is based on the cut off value of risk of propagation of disease to less than 1% of population. This model can also be used as a base to arrive at the 95% CI or margin of error of the diagnostic sensitivity of other infectious diseases with known R_0_ value or based on a presumptive R_0_ value and % prevalence of the disease.

A random population of varying sizes need to be tested to find out the best lower bound of 95% CI or margin of error of the diagnostic test (Russek-Cohen et al., 2011). The width of the 95% CI or margin of error based on a random sample will vary randomly and the 95% CI or margin of error may not always give an idea of the population parameter (Hazra, 2017). However, diagnostic tests intended for the diagnosis of highly infectious diseases with high risk of propagation such as COVID-19 need stringent regulatory standards for pre- and post-market approval.

The negative correlation between the margin of error value of diagnostic sensitivity and number of approved COVID-19 kits suggestes that margin of error or 95% CI of the diagnostic sensitivity is considered by that the regulatory agencies while approving the test kits. Though majority of approved COVID-19 test kits fell under less than 5% margin of error of sensitivity category, several approved kits with margin of error range 5% to 40% were also found and clearly suggests the lack of regulatory standards on the margin of error or 95% CI of diagnostic sensitivity/specificity of COVID-19 test kits.

The regulatory approval of a diagnostic test intended for highly infectious disease such as COVID-19 should not only based on the sensitivity and specificity criteria, but also be based on the 95% CI or the margin of error as a diagnostic test with perfect sensitivity and wider 95% CI or high margin of error could lead to lot of false negative results which in turn facilitate the rapid spread of the diseases.

## 5. Conclusion

A mathematical model had been developed to estimate the mean % secondary infections based on the margin of error of diagnostic sensitivity, % prevalence and R_0_ value. This model recommends a diagnostic test with diagnostic sensitivity ≥ 96% and at least 92% lower bound limit of the 95% CI or ≤ 4% margin of error for highly infectious diseases like COVID-19. Positive correlation was found between the estimated mean % of secondary infection and margin of error and increase in margin of error of a diagnostic test would lead to rapid disease transmission due to false negative cases. Negative correlation was found between the number of COVID-19 diagnostic kits approved by regulatory agencies and margin of error of the test kits suggesting the chance of regulatory approval for a COVID-19 kits is higher when the margin of error is low. Though it is evident that margin of error or 95% CI plays a role in regulatory approval of a COVID-19 test kits, the norms and mathematical models followed by the regulatory agencies were not available. For COVID-19 like highly infectious disease with high risk of propagation certainly need specific regulatory standards on margin of error or 95% CI of the diagnostic sensitivity to curb the transmission of the disease due to false negative cases.

## Author contributions

**Azhahianambi, P:** Conceptualization, Formula derivation, R-software programming, statistical analysis, interpretation of results and drafting the manuscript. **Karthik, K:** Data collection and compilation, data curation and formatting. **Aravindh Babu, RP:** Data curation, supplementary files, manuscript review. **Senthil kumar, TMA:** Data curation, manuscript review and funding. **Balachandran, C and Dhinakar Raj, G:** Conceptualization, funding, manuscript review

## Supporting information

Mathematical model data

Data from finddx

## Data Availability

Data pertaining to the mathematic modelling were provided as supplementary files

## Acknowledgment

The funding received from Department of Biotechnology, Govt. of India for the project entitled Translational Research Planform for Veterinary Biologicals-Phase III (BT/TRPVB/TANUVAS/2011/Phase III dated 17/09/2020) is duly acknowledged

## Declaration on conflict of Interest

The authors of this manuscript declare no conflict of interest

## Notes

### Competing Interest Statement

The authors have declared no competing interest.

### Author Declarations

This study is neither a clinical nor an experimental animal study. Hence dose not require any kind of approval

