## Supplementary material for "A mathematical model to estimate percentage secondary infections from margin of error of diagnostic sensitivity: Useful tool for regulatory agencies to assess the risk of propagation due to false negative outcome of diagnostics": Tables

**Table 1.** Estimated mean % secondary infection at various diagnostic sensitivity and margin of error values where R_0_ is 2.0 and relationship between margin of error and estimated mean % secondary infection

| **95% CI** | **Margin of error** | **Estimated mean % secondary infection** | **Standard Error of the mean** | **Correlation Coefficient (R) and Coefficient of determination (R^2^)** |
| --- | --- | --- | --- | --- |
| ***Diagnostic Sensitivity 99%-100%*** | | | | |
| **98-100** | **1** | **0.2** | 0.14 | **R=1.0**  *P<*0.00001  **R^2^= 1.0**  *p*<0.00001 |
| **96-100** | **2** | **0.4** | 0.16 |  |
| **94-100** | **3** | **0.6** | 0.18 |  |
| **92-100** | **4** | **0.8** | 0.19 |  |
| **90-100** | **5** | **1** | 0.21 |  |
| **88-100** | **6** | **1.2** | 0.23 |  |
| **86-100** | **7** | **1.4** | 0.24 |  |
| **84-100** | **8** | **1.6** | 0.25 |  |
| **82-100** | **9** | **1.8** | 0.27 |  |
| **80-100** | **10** | **2** | 0.26 |  |
| ***Diagnostic Sensitivity 98%*** | | | | |
| **97-99** | **1** | **0.4** | 0.10 | **R=0.99**  *P<*0.00001  **R^2^= 0.99**  *p*<0.0001 |
| **96-100** | **2** | **0.4** | 0.16 |  |
| **94-100** | **3** | **0.6** | 0.18 |  |
| **92-100** | **4** | **0.8** | 0.19 |  |
| **90-100** | **5** | **1** | 0.21 |  |
| **88-100** | **6** | **1.2** | 0.23 |  |
| **86-100** | **7** | **1.4** | 0.24 |  |
| **84-100** | **8** | **1.6** | 0.25 |  |
| **82-100** | **9** | **1.8** | 0.27 |  |
| **80-100** | **10** | **2** | 0.26 |  |
| ***Diagnostic Sensitivity 97%*** | | | | |
| **96-98** | **1** | **0.6** | 0.08 | **R=0.98**  *P<*0.00001  **R^2^= 0.96**  *p*<0.0001 |
| **95-99** | **2** | **0.6** | 0.13 |  |
| **94-100** | **3** | **0.6** | 0.18 |  |
| **92-100** | **4** | **0.8** | 0.19 |  |
| **90-100** | **5** | **1** | 0.21 |  |
| **88-100** | **6** | **1.2** | 0.23 |  |
| **86-100** | **7** | **1.4** | 0.24 |  |
| **84-100** | **8** | **1.6** | 0.25 |  |
| **82-100** | **9** | **1.8** | 0.27 |  |
| **80-100** | **10** | **2** | 0.26 |  |
| **Diagnostic Sensitivity 96%** | | | | |
| **95-97** | 1 | **0.8** | 0.07 | **R=0.95**  *p*=0. 00003  **R^2^= 0.91**  *p*<0.0001 |
| **94-98** | 2 | **0.8** | 0.11 |  |
| **93-99** | **3** | **0.8** | 0.15 |  |
| **92-100** | **4** | **0.8** | 0.19 |  |
| **90-100** | **5** | **1** | 0.21 |  |
| **88-100** | **6** | **1.2** | 0.23 |  |
| **86-100** | **7** | **1.4** | 0.24 |  |
| **84-100** | **8** | **1.6** | 0.25 |  |
| **82-100** | **9** | **1.8** | 0.27 |  |
| **80-100** | **10** | **2** | 0.26 |  |
| ***Diagnostic Sensitivity 95%*** | | | | |
| **94-96** | **1** | **1** | 0.14 | **R=0.94**  *p*=0.00005  **R^2^= 0.88**  *p*<0.0001 |
| **93-97** | **2** | **1** | 0.16 |  |
| **92-98** | **3** | **1** | 0.18 |  |
| **91-99** | **4** | **1** | 0.19 |  |
| **90-100** | **5** | **1.1** | 0.19 |  |
| **88-100** | **6** | **1.3** | 0.21 |  |
| **86-100** | **7** | **1.5** | 0.22 |  |
| **84-100** | **8** | **1.7** | 0.24 |  |
| **82-100** | **9** | **1.9** | 0.25 |  |
| **80-100** | **10** | **2.1** | 0.26 |  |
| ***Diagnostic Sensitivity 90%*** | | | | |
| **89-91** | **1** | **2** | 0.14 | **R=0.52**^NS^  *P*=0.123  **R^2^= 0.27**^NS^  *p*=0.122 |
| **88-92** | **2** | **2** | 0.16 |  |
| **87-93** | **3** | **2** | 0.18 |  |
| **86-94** | **4** | **2** | 0.19 |  |
| **85-95** | **5** | **2** | 0.21 |  |
| **84-96** | **6** | **2** | 0.22 |  |
| **83-97** | **7** | **2** | 0.24 |  |
| **82-98** | **8** | **2** | 0.25 |  |
| **81-99** | **9** | **2** | 0.27 |  |
| **80-100** | **10** | **2.1** | 0.26 |  |

**NS**: Not significant at 5% and 10% level

*****Sensitivity, 95% CI range and margin of error with less than 1% mean secondary infection is highlighted by shading

**Table 2.** Approved COVID-19 diagnostic kits with diagnostic sensitivity and specificity greater than 90% listed in finddx.com

| **Sensitivity >90% (Specificity varies)** | | | | | | | | | | |
| --- | --- | --- | --- | --- | --- | --- | --- | --- | --- | --- |
| **S.No** | **Test platform** | **Number of Total kits listed** | **No. of kits with sensitivity > 90%** | **% of kits with sensitivity > 90%** | **No. of kits with margin of error** | | | | | |
|  |  |  |  |  | **<5** | **5-10** | **10.1-20** | **20.1-30** | **30.1-40** | **40.1-50** |
| **1** | **NAAT** | 41 | 29* | 70.7 % | 10 | 4 | 6 | 4 | 1 | 0 |
| **2** | **Antibody test** | 32 | 13 | 40.6 % | 7 | 5 | 1 | 0 | 0 | 0 |
| **3** | **Antigen test** | 5 | 2 | 40.0 % | 0 | 2 | 0 | 0 | 0 | 0 |
| **Specificity >90% (Sensitivity varies)** | | | | | | | | | | |
| **S.No** | **Test platform** | **Number of Total kits available at finddx.com** | **No. of kits with sensitivity > 90%** | **% of kits with sensitivity > 90%** | **No. of kits with margin of error** | | | | | |
|  |  |  |  |  | **<5** | **5-10** | **10.1-20** | **20.1-30** | **30.1-40** | **40.1-50** |
| **1** | **NAAT** | 41 | 29* | 70.7 % | 11 | 3 | 2 | 0 | 2 | 7 |
| **2** | **Antibody test** | 32 | 13 | 40.6 % | 10 | 2 | 0 | 1 | 0 | 0 |
| **3** | **Antigen test** | 5 | 1 | 40.0 % | 2 | 0 | 0 | 0 | 0 | 0 |

**4 kits have margin of error greater than 50.0*

**Table 3.** Margin of error of NAAT based and antibody-based COVID-19 test kits with regulatory approval and relationship between margin of error and number of regulatory approved COVID-19 test kits

| ***Diagnostic sensitivity > 90%** | | | | | | | |
| --- | --- | --- | --- | --- | --- | --- | --- |
|  | ***M* <5** | ***M*= 5-10** | ***M*= 10.1-20** | ***M*= 20.1-30** | ***M*= 30.1-40** | ***R** | ****R^2^** |
| **Number of approved NAAT based tests** | 10 | 4 | 6 | 4 | 1 | **-0.92**  (*p*=0.023) | **0.85**  (*p*=0.023) |
| **Number of approved Antibody based tests** | 7 | 5 | 1 | 0 | 0 |  |  |
| **Total** | **17** | **9** | **7** | **4** | **1** |  |  |
| **Diagnostic specificity > 90%** | | | | | | | |
|  | ***M* <5** | ***M*= 5-10** | ***M*= 10.1-20** | ***M*= 20.1-30** | ***M*= 30.1-40** | ***R** | ****R^2^** |
| **Number of approved NAAT based tests** | 11 | 3 | 2 | 0 | 2 | **-0.70**^NS^  (*p*=0.188) | **0.49**^NS^  (*p*=0.186) |
| **Number of approved Antibody based tests** | 10 | 2 | 0 | 1 | 0 |  |  |
| **Total** | **21** | **5** | **2** | **1** | **2** |  |  |

***** *Pearson correlation coefficient (R) between the midpoints of margin of error and number of approved kits in each range of margin of error*

****** *Coefficient of determination (R^2^) between the midpoints of margin of error and number of approved kits in each range of margin of error*

**NS**: *Not significant at 5% and 10% level*
